# Optimization of a Novel Automated, Low Cost, Three-Dimensional Photogrammetry System (PHACE)

**DOI:** 10.1101/2023.04.21.23288659

**Authors:** Josiah K. To, Jenny N. Wang, Anderson N. Vu, Lilangi S. Ediriwickrema, Andrew W. Browne

## Abstract

**Introduction:** Clinical tools are neither standardized nor ubiquitous to monitor volumetric or morphological changes in the periorbital region and ocular adnexa due to pathology such as oculofacial trauma, thyroid eye disease, and the natural aging process. We have developed a low-cost, three dimensionally printed **PH**otogrammetry for **A**utomated **C**ar**E** (PHACE) system to evaluate three-dimensional (3D) measurements of periocular and adnexal tissue.

**Methods:** The PHACE system uses two Google Pixel 3 smartphones attached to automatic rotating platforms to image a subject’s face through a cutout board patterned with registration marks. Photographs of faces were taken from many perspectives by the cameras placed on the rotating platform. Faces were imaged with and without 3D printed hemispheric phantom lesions (black domes) affixed on the forehead above the brow. Images were rendered into 3D models in Metashape (Agisoft, St. Petersburg, Russia) and then processed and analyzed in CloudCompare (CC) and Autodesk’s Meshmixer. The 3D printed hemispheres affixed to the face were then quantified within Meshmixer and compared to their known volumes. Finally, we compared digital exophthalmometry measurements with results from a standard Hertel exophthalmometer in a subject with and without an orbital prosthesis.

**Results:** Quantification of 3D printed phantom volumes using optimized stereophotogrammetry demonstrated a 2.5% error for a 244μL phantom, and 7.6% error for a 27.5μL phantom. Digital exophthalmometry measurements differed by 0.72mm from a standard exophthalmometer.

**Conclusion:** We demonstrated an optimized workflow using our custom apparatus to analyze and quantify oculofacial volumetric and dimensions changes with a resolution of 244μL. This apparatus is a low-cost tool that can be used in clinical settings to objectively monitor volumetric and morphological changes in periorbital anatomy.

## Introduction

Photogrammetry is the science of calculating spatial and geometric information from objects based on their photographs.^1^ This technology was traditionally used in the fields of cartography and geodesy in conjunction with the advent of photography in the 19^th^ century. Modern improvements in the portability, resolution, cost of image capture, and advancements in the sophistication of computer software have led to adoption of photogrammetry across many disciplines.^2,3^ In medicine, photogrammetry can offer objective measurements of body anthropometry that may be useful in assessing growth and development, treatment response, and surgical outcomes.^4-7^

In ophthalmology, two-dimensional (2D) photographs have long been used for assessment of clinical and surgical outcomes. Examples of 2D photography in ophthalmology include pre- and post-operative comparisons in oculoplastic surgery, monitoring of the anterior segment angle anatomy, and evaluation of optic nerve fiber anatomy.^8-11^ Gradual introduction of three-dimensional (3D) external anatomy imaging technologies over the past decade achieve shorter acquisition times, higher safety, cost-effectiveness, and greater ease of use.^12,13^

The two most common classes of 3D facial surface imaging technologies include structured light technology and stereophotogrammetry. Structured light scanners are considered “active” because they emit grid patterns of visible or infrared light over an object’s surface. 3D shapes are generated by analyzing and calculating distortions in the projected grid as it optically conforms to the scanned object’s contour.^14^ Stereophotogrammetry scanners are considered “passive” because imaging is performed without projecting light patterns onto the scanned object. Photographs of an object are taken from multiple angles to acquire different surface perspectives, which are used to calculate relative spatial coordinates and geometry in 3D space.^13-15^

Hybrid scanners that utilize both structured light and stereophotogrammetry (i.e., active stereophotogrammetry) have become available for medical use in recent years. The suite of 3dMD products (3dMD, Atlanta, GA) are an example of large, multi-camera setups that simultaneously capture images from multiple angles and additionally use structured light. A number of studies have independently verified their high degree of accuracy and reliability.^16-19^ However, due to the high cost, limited portability, and need for frequent recalibration of these 3dMD products, a number of companies have developed handheld, single lens reflex device alternatives such as the Vectra H2 (Canfield Scientific Inc., Fairfield, NJ) and Artec Eva (Artec, Luxembourg).^20,21^ These devices benefit from a high degree of portability and lower cost at the expense of accuracy given the sequential capture method. Despite the decreased accuracy, comparison studies have shown sufficient accuracy for use in many clinical applications.^12, 21-24^ Still, these handheld devices cost thousands of dollars, which may preclude their widespread adoption in general clinical practice. To address the cost concern, Rudy et al. demonstrated the feasibility of using the iPhone X (Apple, Inc., Cupertino, CA) for 3D facial capture in the setting of plastic surgery.^25^ A breakdown of costs, 3D scanning method, and relative accuracies for each of the available devices on the market are summarized in Table 1.

**Table 1.** Stereophotogrammetry devices currently available on the market.^26-29^

| Product Name | Cost | 3D Scanning Method | Relative Accuracy |
| --- | --- | --- | --- |
| 3dMD face | \$10000 | Active stereophotogrammetry | 0.2mm |
| Vectra H2 | \$8000 | Stereophotogrammetry | 1.2mm |
| Artec Eva | \$19800 | Structured-light 3D scanner | 0.1mm |
| iPhone X (refurb) | \$200 | Active stereophotogrammetry | 1mm |

3D imaging devices, including the 3dMD and Vectra M3 systems are suitable for characterizing the periocular region.^30-33^ However, to the best of our knowledge, no studies have investigated the optimization and use of low-cost photogrammetry systems for periocular applications. The purpose of this study was to optimize and evaluate a low-cost (<$300 USD), 3D printed photogrammetry acquisition system for quantitative analysis of periorbital and ocular adnexa morphology and volumetry.

## Methods

### Study Design

This study received Institutional Review Board approval from the University of California, Irvine and was conducted in accordance with the Declaration of Helsinki. Due to limitations of the approved IRB, only portions of faces are shown. Studies performed were HIPAA-compliant and all enrolled participants provided written informed consent.

In this study, a novel data capture system and manual analysis protocol called PHotogrammetry for Automated CarE (PHACE) was evaluated for dimensional accuracy and precision by comparing 3D rendered facial models to real world measurements. This study included 15 healthy adults between the ages of 20 and 65 years (mean age 40 ± 20 years). There were 9 males and 6 females with Fitzpatrick skin types 2, 4, and 6. We evaluated how the number of images from different angles impact 3D model quality when models were generated using several different photogrammetry software tools. We further assessed the precision and accuracy of rendered models by performing depth and volumetric analysis of phantom lesions placed on human subjects. Lastly, the PHACE system was evaluated in an anophthalmic male between 35-40 year-old with Fitzpatrick skin type 2 with and without his left ocular prosthetic.

### Imaging System

The imaging system utilizes two off-the-shelf motorized turntables modified with custom designed 3D printed phone mounts 1.5ft from the platform base and 1.25ft from each other. Each rotation device was clamped onto height adjustable stands with illumination ring lights. Each ring light was set to a brightness of 24,000 lux and 4400K color temperature.

Images were acquired using two Google Pixel 3 smartphones (Android 11 operating system) using the Manual Camera app (Lenses Inc.). User-defined camera settings included: ISO 55, shutter speed 1/80 seconds, focal distance 0.46m (1.5ft), 8MP resolution, portrait orientation lock, and automatic repeating shutter at an acquisition rate of 1 photo/second.

Each smartphone camera was positioned 45cm from the subjects’ glabella with the rotational axis between phones being 38cm apart at the base. The minimum and maximum vergence angles were 0° and 140.8°. The axis of rotation for each smartphone-rotation device was centered on the face. Subjects placed their faces through a foam board cutout with checker patterns for image registration (Figure 1-A). Subjects were instructed to remain motionless with a relaxed facial expression and closed eyes. Pictures of the phantom lesions and the anophthalmic human subject were taken at a rate of one photograph/second and a total of 90 images were processed per subject.

**Figure 1.**
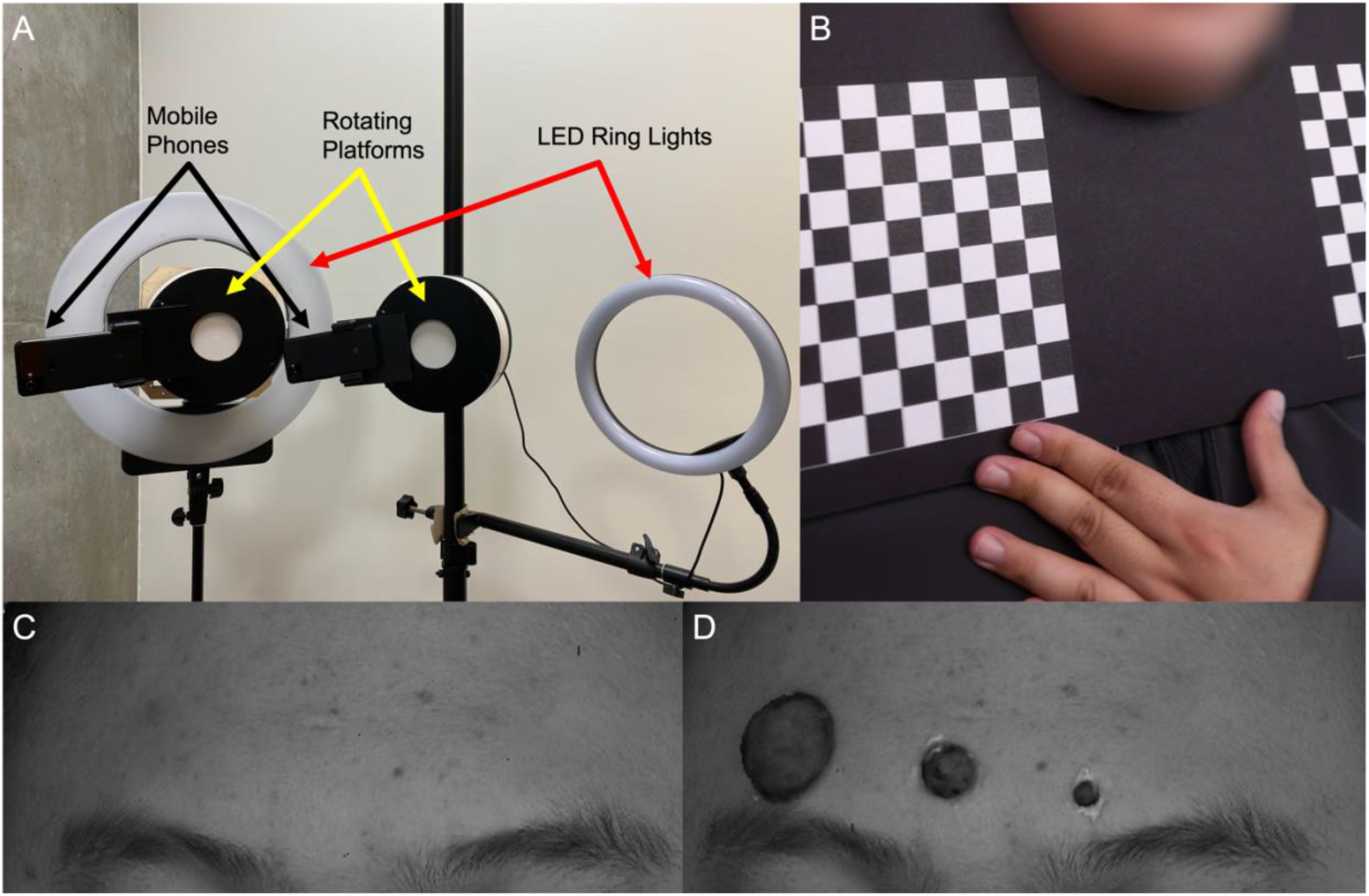
A) Photogrammetry apparatus with two smartphones attached to rotating platforms illuminated by 2 ring LED lights. B) Subjects place their faces through a backdrop with reference checkered squares. Rendered faces without domes (C) and with domes (D). ^*\**^*Pictures have been modified to adhere to Medrxiv picture policy. Please contact corresponding author to request access to this material*.

### Generating 3D Models

Photographs were rendered as 3D facial models and analyzed on a Gigabyte Z390 Aorus Ultra Gaming PC running Windows 10 with an Intel Core i9-9900k 8-core CPU @ 3.6GHz, 48GB of RAM, and a Nvidia GeForce RTX 2080 graphics card. After photographs were taken, a custom python script split RGB images into red, green, and blue channels. Blue channel photos were then imported into four photogrammetry software programs and the quality of rendered models was evaluated using Autodesk’s 3D modeling software.

The four different photogrammetry software tools evaluated were Metashape (Agisoft, St. Petersburg, Russia), Meshroom (AliceVision, Open-source), Pix4Dcloud (Pix4D S.A., Prilly Switzerland), and Zephyr3D (3DFLOW, Verona, Italy). The data set used for comparing photogrammetry software utilized the same set of 90 photographs. Each software was set to the highest settings for alignment, point cloud generation, and mesh rendering. Models from each photogrammetry software were exported as a wavefront object file (.obj file) and then imported into Autodesk’s Meshmixer 3D modeling software for scaling and comparison analysis. The overall quality of rendered models from each photogrammetry software was visually evaluated. Accuracy and precision were qualitatively evaluated by assessing how similar 3D models resembled the human subject face using two metrics: facial shape and surface texture (smooth or rough).

The relationship between the number of images required to be processed without a loss of model precision and the required processing time was determined by both visually and quantitatively assessing model topographical variance. Datasets for modeling three human subjects were imported into Metashape where models were rendered using 120, 100, 90, 80, 60, 40, 30, 20, and 10 photos and their processing times were recorded. Each model was exported as a ‘.obj’ file into Meshmixer, scaled, and then exported into CloudCompare (CC), an open-source point cloud software, for displacement analysis. In CC, the facial models were aligned, cropped, and registered using the Iterative Closest Point (ICP) function. The models constructed from 120 photos were used as a reference to compare models rendered models smaller subsets of the 120 source photograph dataset for each subject. Model quality was both quantitatively and qualitatively evaluated by evaluating smoothness of the topographical appearance (qualitative) and and by analyzing the distribution of mesh face displacement compared to the reference models (quantitative). Models with greater variance compared to reference models were deemed lower quality.

### Volumetric Analysis of Phantom Lesions

To quantify the volumetric accuracy of the PHACE system, three 3D printed hemispheric phantom lesions of different sizes were affixed above the brow line to simulate facial lesions with known volumes. The phantom lesions were printed on a Prusa i3 MKS (Prusa Research, Prague, Czech Republic) 3D printer using Polylactic Acid (PLA) filament (Hatchbox, California, USA) with a layer resolution of 150μm. After printing, the surface texture was roughened using 2000 grit sandpaper to reduce surface reflectivity. The volumes of the small, medium, and large phantom lesions were calculated using the volume formula for a hemisphere, 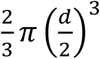, where *d* was the diameter of the 3D printed hemisphere measured with a caliper at a resolution of 0.01mm (Mitutoyo digital caliper). Models of a face generated from images acquired with and without hemispheres attached to the face were exported from Metashape to CC where each model was manually aligned, and models were registered to each other. The paired models were then imported into Meshmixer where the models without hemispheres were Boolean subtracted from the models with phantom lesions. Each phantom lesion’s volume was measured using Meshmixer’s analysis stability tool and then compared with the manually calculated volume.

### Morphological Analysis in Anophthalmic Human Subject

The PHACE system was further validated by comparing the depth displacement of the rendered model of the left orbit of an anophthalmic human subject to real world measurements using a Hertel exophthalmometer. The anophthalmic human subject was a male between 35-40 years-old with Fitzpatrick skin type 2 and a left ocular prosthetic. Models of the subject with and without the ocular prosthetic were rendered in Metashape, imported into CC, and registered to each other. A color map was calculated using orbital depth differences between mesh models with and without the ophthalmic prosthetic. The maximum depth displacement between the rendered facial models with and without the ocular prosthesis was compared to measurements made using the manual exophthalmometer.

## Results

We compared models generated from four different photogrammetry software programs (Figure 2): Metashape (Agisoft, St. Petersburg, Russia), Meshroom (AliceVision), Pix4D (Prilly, Switzerland), and Zephyr3DLite (Verona, Italy) utilizing the highest setting for each software. Models generated using Metashape (Figure 2-A) qualitatively had the most facial normal-appearing facial features while minimizing excessive mesh surfaces that cause unwanted topographical textures or obscure subtle facial features (Figure 2 B-D).

**Figure 2.**
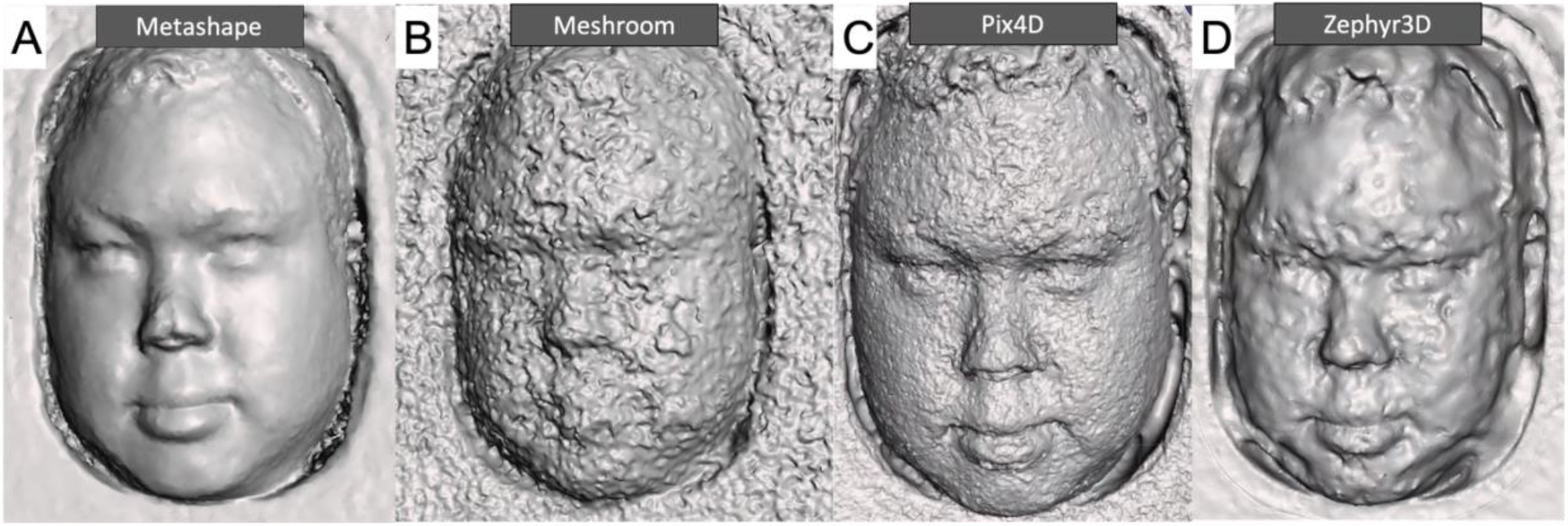
Comparison of four photogrammetry software packages to render facial models. The same photography dataset was used for Agisoft (A), Meshroom (B), Pix4D (C), and Zephyr3D (D).

Next, subsets of 10 to 120 photos were processed and compared using 3D mesh models in CC. It was found that at least 90 photos (25-minute processing time) were required to create a 3D model without compromising precision (Figure 3A – B). The facial model from 120 photos varied from -1.92mm to 2.36mm compared to the mean facial surface. The model in Figure 3D used 20 photos and varied -4.01mm to 2.48mm.

**Figure 3.**
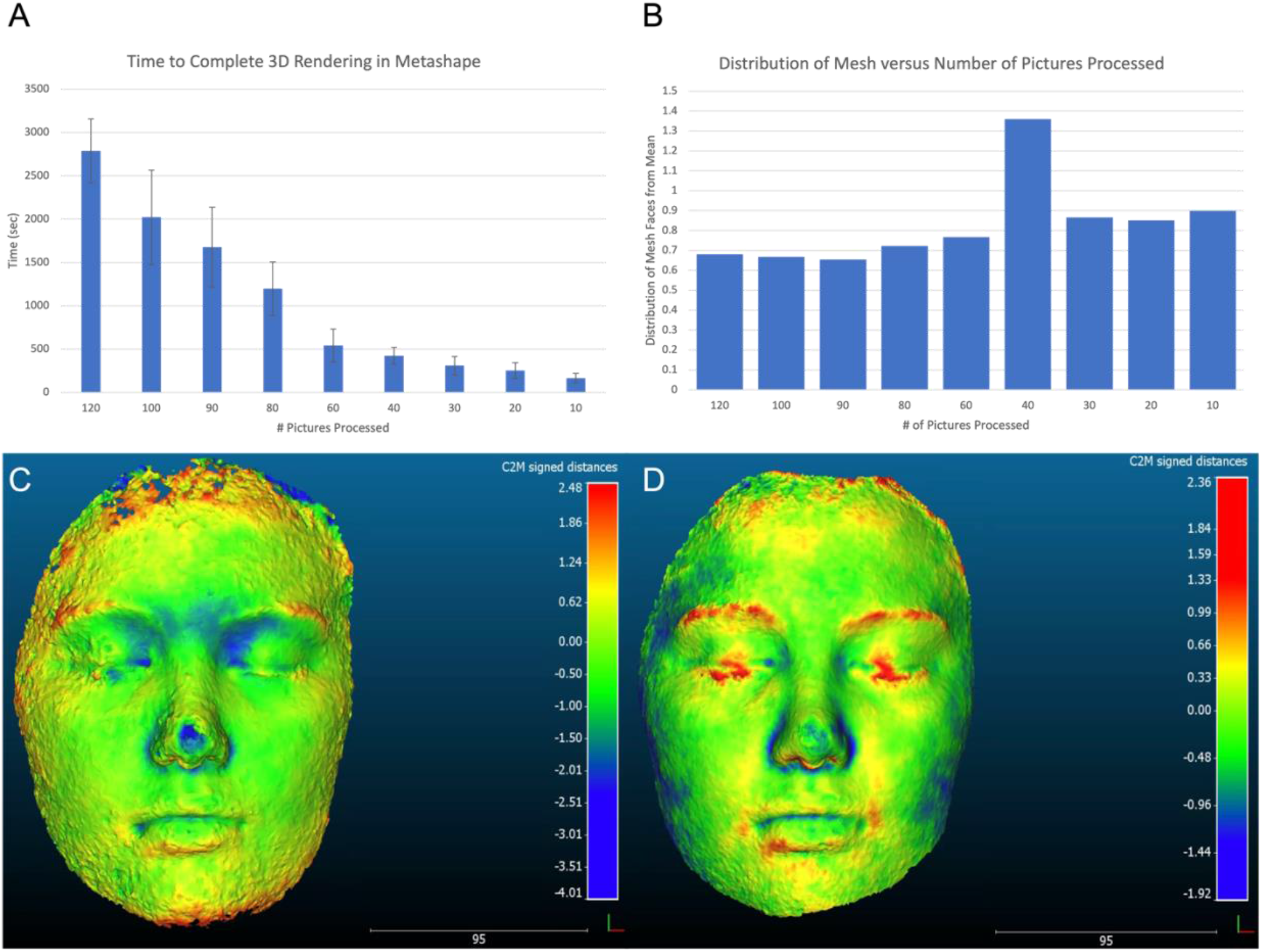
Average distribution of the mesh faces from the mean when using 10 to 120 pictures (A). Average time to create a 3D model using 10 to 120 photos (B). Models generated using 20 (C) and 120 photographs (D).

The hemispheric phantom lesion volumes measured using Meshmixer to evaluate models produced by the PHACE system are shown in Table 2. The PHACE system yielded a 2.5% volume error (52.4μL) compared with the manually calculated reference volume of 2060μL for the large 3D printed hemispherical phantom. A similar error of 2.5% was found for the medium sized (244μL) phantom. The smallest phantom demonstrated the greatest absolute error of 7.6% which accounted for a volumetric difference of 2.1μL.

**Table 2.**
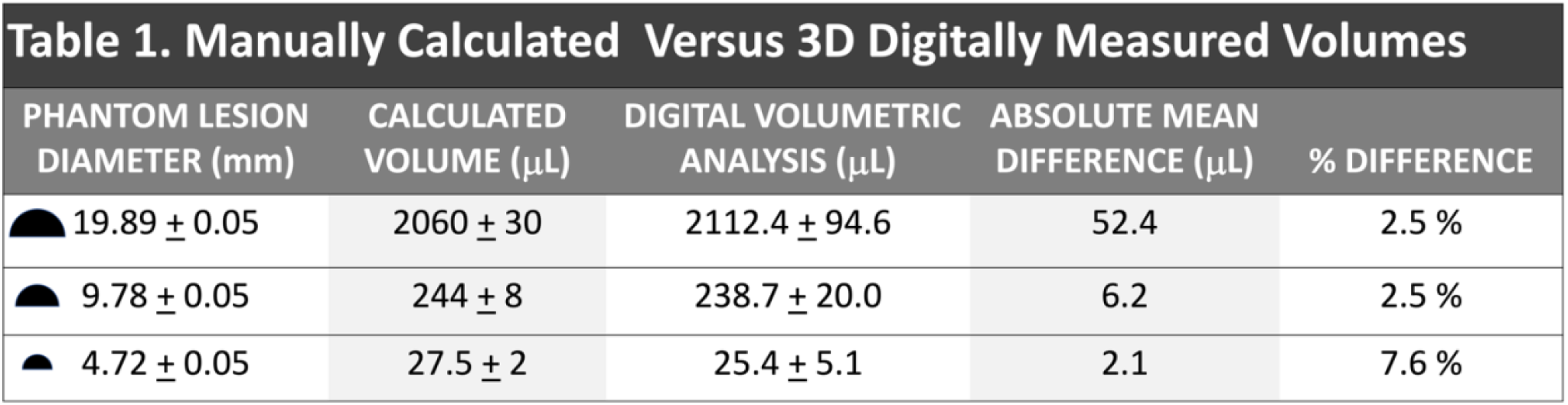
Manually calculated versus 3D digitally measured volume for the three phantom lesion sizes.

| Table 1. Manually Calculated Versus 3D Digitally Measured Volumes |  |  |  |  |
| --- | --- | --- | --- | --- |
| PHANTOM LESION<br>DIAMETER (mm) | CALCULATED<br>VOLUME (μL) | DIGITAL VOLUMETRIC<br>ANALYSIS (μL) | ABSOLUTE MEAN<br>DIFFERENCE (μL) | % DIFFERENCE |
| 19.89 ± 0.05 | 2060 ± 30 | 2112.4 ± 94.6 | 52.4 | 2.5 % |
| 9.78 ± 0.05 | 244 ± 8 | 238.7 ± 20.0 | 6.2 | 2.5 % |
| 4.72 ± 0.05 | 27.5 ± 2 | 25.4 ± 5.1 | 2.1 | 7.6 % |

In the anophthalmic subject (Figure 4), the left eye depth without the ophthalmic prosthetic was measured with a Hertel exophthalmometer. The exophthalmometer quantified a difference 3.5mm of lid protrusion with and without the prosthetic in place. PHACE measured a depth difference of 4.22mm (Figure 4).

**Figure 4.**
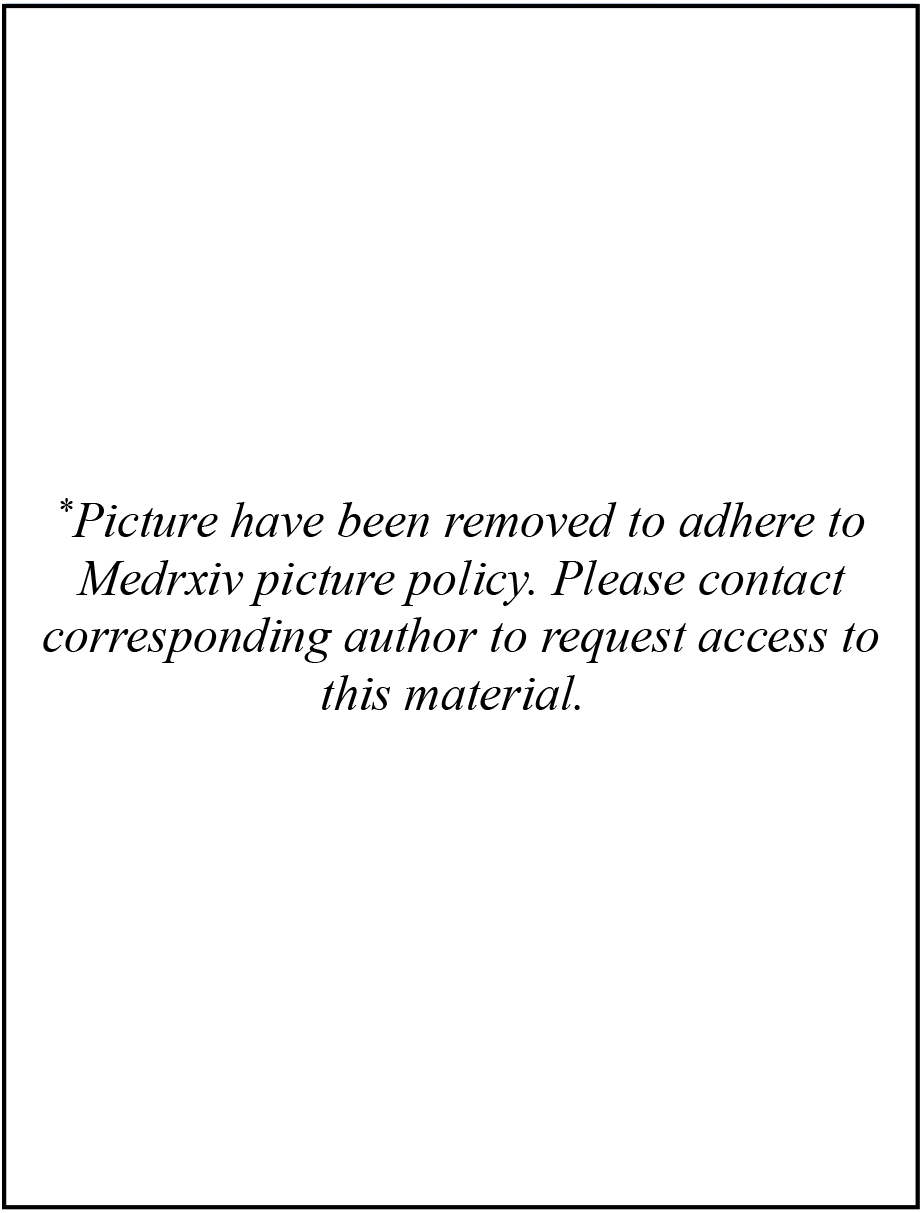
Model depth analysis of human subject with ophthalmic prosthesis (top), without prosthesis (middle), and colorized depth map (bottom) representing differences in depth. The automated photogrammetry model with overlaying depth map shows a depth difference of -4.22 mm.

## Discussion

3D anthropometry is a continuously evolving field as technology continues to improve. There are few studies and specific technologies that evaluate 3D periorbital anthropometry for clinical use. In this study, we optimized our PHACE system with focus on optimizing both low-cost components while maintaining topographical resolution to computationally reconstruct human faces for digital quantitative analysis. This study used free software and a novel volumetric methodology for depth analysis to characterize the PHACE system.

### Optimized Automated Stereophotogrammetry

The PHACE system was developed with stereophotogrammetry at its core. Unlike high and mid-cost structured light scanning technologies, photogrammetry is the most ubiquitous, economical and versatile 3D modeling technique currently available. Any low-cost camera system (e.g., a smartphone or raspberry pi camera) in combination with an affordable or open-source photogrammetry software can be used to safely and accurately render high quality models with potential utility in healthcare.

Although Murta et al. demonstrated the utility and clinical need for quantitative 3D methods of evaluating facial morphology in the setting of ocular disease/ophthalmic interventions (e.g., orbit decompression for thyroid eye disease) they use an expensive stereophotogrammetric system costing thousands of dollars.^34^ There are no well-established or routinely employed affordable automated systems that produce high quality models for morphologic and volumetric evaluation in ophthalmology. The PHACE system uses off-the-shelf and 3D printed components to create an automated imaging system to reproducibly procure photographs of subject faces from multiple different angles with very little human input. Reconstructing 3D facial models for medical evaluation requires clear, high-resolution photographs to be able to discriminate millimeter and sub-millimeter sized features. Automated camera actuation and photograph acquisition remove interoperator variability, which is important in 3D model measuring reproducibility. Ceinos et al. found that there was minimal inter-examiner variability in obtaining facial measurements from stereophotogrammetry scans when scans are acquired manually^35^, and automating the process further reduces potential variability. Therefore, the PHACE system (Figure 1-A) is a clinically optimized, low-cost automated stereo-capture system using off-the-shelf components.

The photogrammetry software is equally important to the 3D model optimization. Not all photogrammetry software tools perform equally, and Figure 2 compares the quality of rendered models between different photogrammetry software using the same 100 photographs. Metashape reconstructs faces with the highest detail, while also minimizing the extraneous mesh noise (Figure 2A). In contrast, the model constructed by Pix4D exhibits detailed facial features (i.e., the contour of the eyelids and adnexa can be distinguished); however, the model has significant mesh noise that show an artificial surface texture and can obscure subtle surface detail (Figure 2C). Guo et al. established several landmarks that are useful in the analysis of periorbital anthropometric scans, including the medial canthus and lateral canthus, and upper and lower eyelid margin.^35^ With significant mesh noise and artificial surface texture in Pix4D reconstructions, such important landmarks are indistinguishable, and thus not useful to track clinical progress over time. Both Meshroom and Zephyr3D create lower quality mesh models that are qualitatively unsuitable for clinical use (Figure 2B and 2D). Therefore, the PHACE protocol adopted Metashape photogrammetry software to reconstruct all facial models.

The final significant factor in optimizing model reconstruction is collecting an optimal number of images. When reducing the number of images processed, the time required to generate images is reduced (Figure 3A). This finding confirms Maas et al. who found that computational effort grows exponentially with the number of photos processed.^36^ However, when reducing the number of images, the precision of the model is also reduced. With fewer photos, there is an increased distribution of mesh faces displaced from the surface of the face (Figure 3B). The comparison between the two sample facial reconstructions (Figure 3C and 3D) highlights the reduced model precision with fewer photos. Ultimately, we found that approximately 90 photos were required to create a 3D model without compromising precision.

### Validation of Automated Stereophotogrammetry

Two methods were used to validate our novel automated photogrammetry system: quantitative volumetric analysis of phantoms and exophthalmometry in a patient with an ocular prosthetic. Prior studies have only evaluated the dimensional accuracy by measuring displacement of rendered models to a reference.^38,39^ However, measuring differences in linear distance between rendered and reference models is limited to interpreting 3D volumetric changes from a 2D imaging plane. Therefore, we quantitatively measured volumes to objectively assess dimensional and morphological accuracy. Our novel methodology to assess digital facial reconstruction techniques used phantom lesions made of 3D printed hemispheres attached to the ocular adnexa to evaluate volumetric and dimensional accuracy. Therefore, we custom designed 3D printed hemispheric phantom to compare known volumes with volumes calculated from PHACE models. The results shown in Table 2 demonstrate that the automated photogrammetry system is able to measure an average volume as small as 244μL^3^ with only a 2.5% difference. Because the double-sided tape attaching the hemispheres to the ocular adnexa is 0.1mm thick, 7.5μL must be subtracted from the artificially increased, digitally measured volume of the hemisphere on the ocular adnexa. Therefore, the photogrammetry system was able to digitally quantify the 244μL hemisphere with an accuracy of an average of 238.7 ± 20μL, which is a 2.5% error and less than the volume of uncertainty due to error propagation from the resolution accuracy of the digital calipers (6.6%). The 27.5μL small hemisphere was reconstructed with an average accuracy of 25.4 ± 5.1μL, which is a 7.6% difference and greater than the error propagation due to the digital caliper resolution. Therefore, the PHACE system can accurately recapitulate small volumes to within 2.5 to 7.6% accuracy. This is more accurate than the published findings of 5-14 mL volume differences when measuring volumetric changes in facial swelling after orthognathic surgery using 3D stereophotogrammetry scans of the head and neck using the 3dMDface stereophotogrammetry system.^40^

Quantitative depth analysis was performed on rendered models from a human subject with and without his prosthetic eye and compared to measurements obtained via a Hertel exophthalmometer (Figure 4). The change in depth digitally measured by the PHACE system was -4.22mm, while the exophthalmometer measured a change in depth of -3.5mm However, the exophthalmometer’s resolution is limited to millimeters and can only be subjectively estimated by in 0.5mm increments. Therefore, comparing digital depth measurements to human manual exophthalmometer measurements can easily suffer from subjective user variability. Experts in the field agree that measurements with the Hertel exophthalmometer are not exact but should be repeatable within 1-2mm.^41^ Therefore, our measurements from the PHACE digital model fall within the standard error limits of the standard exophthalmometer measurement ability.

### Limitations and Future Directions

A significant consideration when using a 3D model reconstruction technique for quantitative change analysis of human faces is that subtle micro-expressions will drastically reduce the comparability between models and ultimately lower the sensitivity of the analysis. In theory, after reconstructed models are registered to each other in 3D space, all changes detected are due to external factors altering the region of interest. In the medical context, changes between models will ideally be due solely to medical conditions altering tissue volume and depth. However, facial micro-expressions as simple as a subtle smile or frown will alter the 3D model. Brons et al. found that involuntary facial expressions can make significant differences in 3D images, particularly along the nasolabial region.^42^ The error from involuntary facial expressions may propagate and amplify during model alignment and registration in 3D space. When reconstructed models are registered in 3D space, subtle changes to any model region can alter the alignment and registration of the models. To address this concern, we asked subjects to close their eyes and relax their faces. The effect of microexpression on open eyelid morphology will need to be investigated thoroughly before systems like PHACE can reliably quantify changes in tissue volumes affecting eyelid morphology.

To procure reliable measurements on 3D rendered faces, visual landmarks consistent across varying human faces needs to be established. Several groups have investigated the breadth of periorbital anthropometric technologies available and have helped define standardized landmarks that can be used, including the medial canthus and lateral canthus, and upper and lower eyelid margins.^36,43,44^ The implementation of standardized landmarks will help align photos more accurately and allow clinicians to make accurate measurements using digitally reconstructed models, and in turn to compare clinical changes over time. Future directions to make this technology more clinically useful including automating MeshMixer in such a manner so that clinicians can readily obtain image output measurements without engaging any lengthy imaging analysis protocols.

## Conclusions

We optimized parameters for an automated stereophotogrammetry imaging system using a photogrammetric software protocol. We have also demonstrated a novel method for evaluating volumetric dimensional accuracy of 3D reconstruction techniques by comparing digitally measured volumes of 3D printed hemispheric phantom lesions against their calculated volumes. The PHACE system can accurately recapitulate volumes as small as 244μL to approximately 2.5%. This affordable and easy-to-use stereophotogrammetry system should be considered in clinical settings to evaluate volumetric dynamics over the course of care.

## Data Availability

All data produced in the present study are available upon reasonable request to the authors

## Notes

Financial Support: Arnold and Mabel Beckman Foundation Fellowship, Research to Prevent Blindness unrestricted grant to UCI Department of Ophthalmology, and a UCI ICTS NIH KL2 Grant number KL2 TR001416.

### Competing Interest Statement

The authors have declared no competing interest.

### Funding Statement

Arnold and Mabel Beckman Foundation Fellowship, Research to Prevent Blindness unrestricted grant to UCI Department of Ophthalmology, and a UCI ICTS NIH KL2 Grant number KL2 TR001416.

### Author Declarations

Ethics committee/IRB of the University of California Irvine gave ethical approval for this work

